# Methylation of the glucocorticoid receptor gene (NR3C1) in dyads mother-child exposed to intimate partner violence in Cameroon: Association with anxiety symptoms

**DOI:** 10.1101/2022.08.14.22278760

**Authors:** Dany Laure Wadji, Naser Morina, Chantal Martin-Soelch, Chantal Wicky

## Abstract

**Background:** The glucocorticoid receptor (GR), which is encoded by the *NR3C1* (Nuclear Receptor Subfamily 3 Group C Member 1) gene plays an important role in the modulation of the hypothalamic-pituitary-adrenal (HPA) axis activity by providing feedback regulation which allows termination of the stress response. Little is known about epigenetic programming at the level of NGFI-A putative binding site (CpG) of the *NR3C1* exon 1F in dyads mother-child exposed to intimate partner violence (IPV) more specifically in an unstudied region such as the Sub-Saharan Africa where levels of violence are very high.

**Objective:** Examine *NR3C1* exon 1F methylation in response to IPV and possible association with cortisol concentration and mental health.

**Method:** We recruited 20 mother–child dyads exposed to IPV and a control group of 20 mother–child dyads not exposed to IPV. We administered self-reported questionnaires to measure mother’s mental health and collected saliva samples for cortisol dosage and bisulfite sequencing of DNA methylation.

**Results:** Regarding the mothers, our results showed a significant difference in methylation level at CpG 16-21 sites of the *NR3C1* exon 1F promoter region between the groups. In the exposed group as compared to the control group, there was a significant positive association between the level of methylation at CpG 16-21 sites and mother’s mental health in particular anxiety symptoms. However, we did not find any significant correlation between methylation level and cortisol concentration. In children, we did not find any significant result.

**Conclusion:** This study highlights a NGFI-A putative binding site (CpG 16-21) that is more methylated in mothers exposed to IPV and which may have the potential to confer vulnerability for psychopathologies.

## Introduction

Approximately 30% of women worldwide are victims of intimate partner violence (IPV) (1). In sub-Saharan Africa, the rate of IPV is particularly high, with an overall prevalence of 36% exceeding the global average, due to cultural habits and cultural norms (2, 3). IPV is associated with a wide range of physical and psychological symptoms for women and their children such as injuries, anxiety, depression, chronic pain or post-traumatic stress disorder (4, 5). The effects of IPV are not limited to poor physical and mental health of direct victims (6, 7) but also have important overwhelming economic burden on society in terms of direct and indirect costs related to health care services, social rehabilitation programs, and adult/juvenile crime (8).

At a physiological level, the hypothalamic-pituitary-adrenal (HPA) axis plays a central role in in the body’s responses to stress and can therefore be central in understanding how adverse environment including IPV is associated with various health issues (9). Upon acute stress, a cascade of events, including the release of hypothalamic corticotropin-releasing factor (CRF), activates the pituitary-adrenal system leading to the production of cortisol in the circulating blood, as well as in body fluids such as saliva (10, 11). Increasing cortisol level will in turn induce a negative feedback on the hypothalamus leading to the inhibition of CRF synthesis and eventually dampening of the HPA axis response to stress (12, 13). In contrast, chronic stress has the potential to dysregulate the HPA axis, resulting in blunted cortisol reactivity, which increases risk of a number of stress-related physical and mental diseases (14, 15).

Previous research suggested an association between IPV and HPA axis activity. For example, high cortisol concentration was observed in women who experienced IPV as compared to a control group, through salivary/plasma cortisol assessment with measurements such as cortisol awakening response (CAR) (16) and area under the curve with respect to ground (AUCg) (17), or through hair cortisol assessment (18, 19). Further evidence suggested a correlation between HPA axis dysregulation as shown by elevated cortisol production and enhanced mental health problems (20–22). However, the molecular mechanisms underlying the impact of IPV on mental health is still poorly understood.

Cortisol induces stress responses by binding to the glucocorticoid receptor (GR), which is encoded by the *NR3C1* (Nuclear Receptor Subfamily 3 Group C Member 1) gene. This receptor plays an important role in the modulation of HPA axis activity by providing feedback regulation which allows termination of the stress response (19, 23, 24). This negative feedback loop which provides a means for HPA axis homeostasis, relies on the availability of glucocorticoid receptors (GR) (25). The activity of the *NR3C1* receptor can be modulated at the transcriptional level of the gene which can be downregulated upon methylation of its promoter region. The *NR3C1* gene consists of nine untranslated alternative spliced exon 1 variants and 8 coding exons (26). The region encompassing exon 1 variants also functions as a proximal promoter and includes multiple methylation-sensitive cytosine phosphate-guanine (CpG) dinucleotide repeats (27). The most studied promoter region is the exon 1F which includes 47 CpG sites as well as several binding sites of the transcription factor NGFI-A (nerve growth factor-inducible protein A) (see Fig. 1). Binding of NGFI-A to exon 1F promoter region increases GR expression, however, increased methylation of *NR3C1* in this region reduces glucocorticoid receptor gene expression, thereby limiting the availability of glucocorticoid receptors (28, 29). Consequently, the negative feedback loop regulating HPA axis activity is disrupted (30, 31) putting people at greater risk of mental health problems (32, 33).

**Fig 1.**
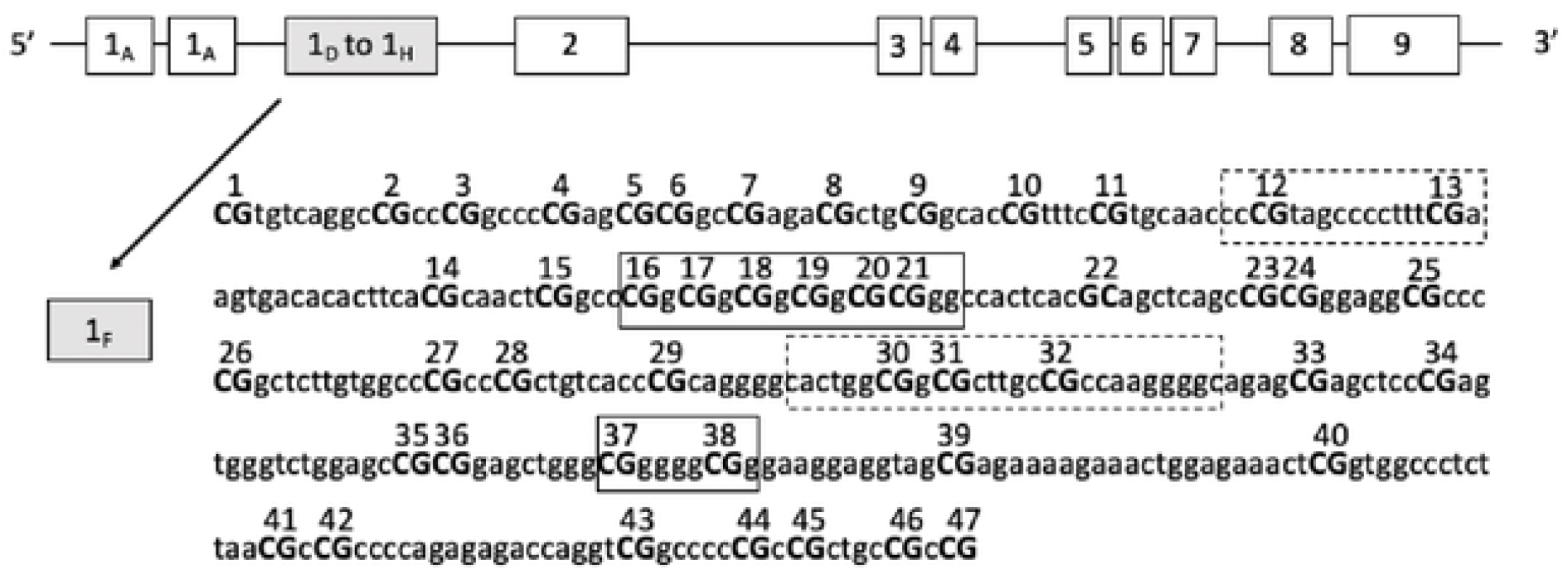
*NR3C1* gene structure. The *NR3C1* gene covers a region of more than 80 kb on chromosome V. It contains 8 coding exons (2-9) and 9 alternative spliced 5’ non coding exon 1 variants. The region encompassing exon 1 variants is also known to function as a proximal promoter. Methylation of CpG dinucleotides (capital letters, 1-47) of exon lF promoter region is examined in this study. Boxes represent known or putative canonical NGFI-A binding sites and broken-line boxes represent non canonical NGFl-a binding sites.

Previous intergenerational studies have showed an association between IPV and DNA methylation of several genes. For example, a correlation between IPV and *NR3C1* DNA methylation in exon 1F promoter region was shown in offspring of mothers, victims of IPV during pregnancy (34). This study was the first intergenerational study on dyads mother-child to indicate that *NR3C1* promoter methylation is associated with IPV. Further, grandmaternal (G1) exposure to interpersonal violence during pregnancy was associated with several differentially methylated genes in grandchildren (G3) (e.g. CFTR, CORIN, BARX1 and SMYD3 genes) (35). However, they did not examine epigenetic modification in mothers (G2). A more recent study examined genome-wide epigenetic modifications in response to domestic violence. It revealed that two genes involved in neuronal development, BDNF (brain-derived neurotrophic factor) and CLPX (caseinolytic mitochondrial matrix peptidase chaperone subunit), were differentially methylated in adolescents and their mothers exposed to domestic violence (36).

Taken together, prior studies with dyads mother-child are pointing towards a role of epigenetic programming in response to IPV at the level of several genes at several CpG sites within their promoter. However, the epigenetic mechanism underlying the link between IPV and poor mental health is still not fully understood. The *NR3C1* exon 1F gene, which appears to be particularly important because of its link to the HPA axis, has received so far very little attention in studies on IPV. In recent works from our group with a larger sample of dyads mother-child exposed to IPV, including the dyads examined in the present paper, we showed on one hand that mothers exposed to IPV compared to the control group have high levels of anxiety and depression symptoms and that their children exhibited high levels of externalizing symptoms such as delinquent and aggressive behaviour (7) and on the other hand exposed mother presented increased HPA-axis activity i.e. higher post-awakening cortisol concentrations (17). Exploring *NR3C1* promoter methylation in the region of exon 1F and possible association with total cortisol concentration and symptoms of psychopathology in this sub-sample, can provide a stronger argument for long-term consequences of IPV particularly in an unstudied region like Sub-Saharan Africa where levels of violence are very high.

This epigenetic study on IPV targeting a traumatized population of women and children, aims to provide evidence of DNA methylation of *NR3C1* exon 1F promoter region. We hypothesized that: 1) the exposed group will present significant higher DNA methylation compared to the control group, 2) global *NR3C1* DNA methylation levels and CpG specific sites DNA methylation level in the promotor region, including known transcription binding factor binding sites, would be significantly associated with cortisol concentration and current symptoms of psychopathology.

## Materials and Methods

### Participants

The study received approval from the National Commission for Ethics in Cameroon (N0 2019/02/1141/CE/CNERSH/SP). We recruited a total of 75 mother-child dyads with the help of the Association for the Fight against Domestic Violence (ALVF) a non-profit organisation based in Cameroon, which aids female victims of IPV. However, the present study was conducted on a subsample of 40 mother-infant dyads because in this pilot study we were eager to see if the epigenetic data collection and their analysis were possible in the Cameroonian context. Thus, we collected 40 samples from 20 mother-child dyads exposed to IPV (mean age±SD: 37.05 ± 7.515, range 25–53 years) and 40 samples from 20 mother-child dyads not exposed to IPV (mean age±SD: 37.65 ± 4.03, range 26–44 years). The recruitment was done with the help of community agents of ALVF who performed a door-to-door sensitization and awareness campaign for our study. Mothers were aware that their participation in the study was voluntary and that not agreeing to participate would have no consequences on service provision by ALVF. The inclusion criteria were (1) being physically, sexually, and/or psychologically abused or injured in the past 12 months; (2) having a biological child aged 2–18 who have seen and heard violent acts, seen injuries resulting from the violence, or been told about the violence in the past 12 months (if a mother had more than one child in this age range, she was asked to select the one whom she felt had been most exposed); and (3) the ability to express herself in French or English. Mothers who did not speak and understand either French or English were excluded, as well as those with children older than 18. All mothers denied any history of medical or neurological illness or use of psychotropic medications. Demographic information was collected, including the participant’s age, sex, level of education, number of children, matrimonial status, profession, ethnic group and religion (see Table 1).

**Table 1.**
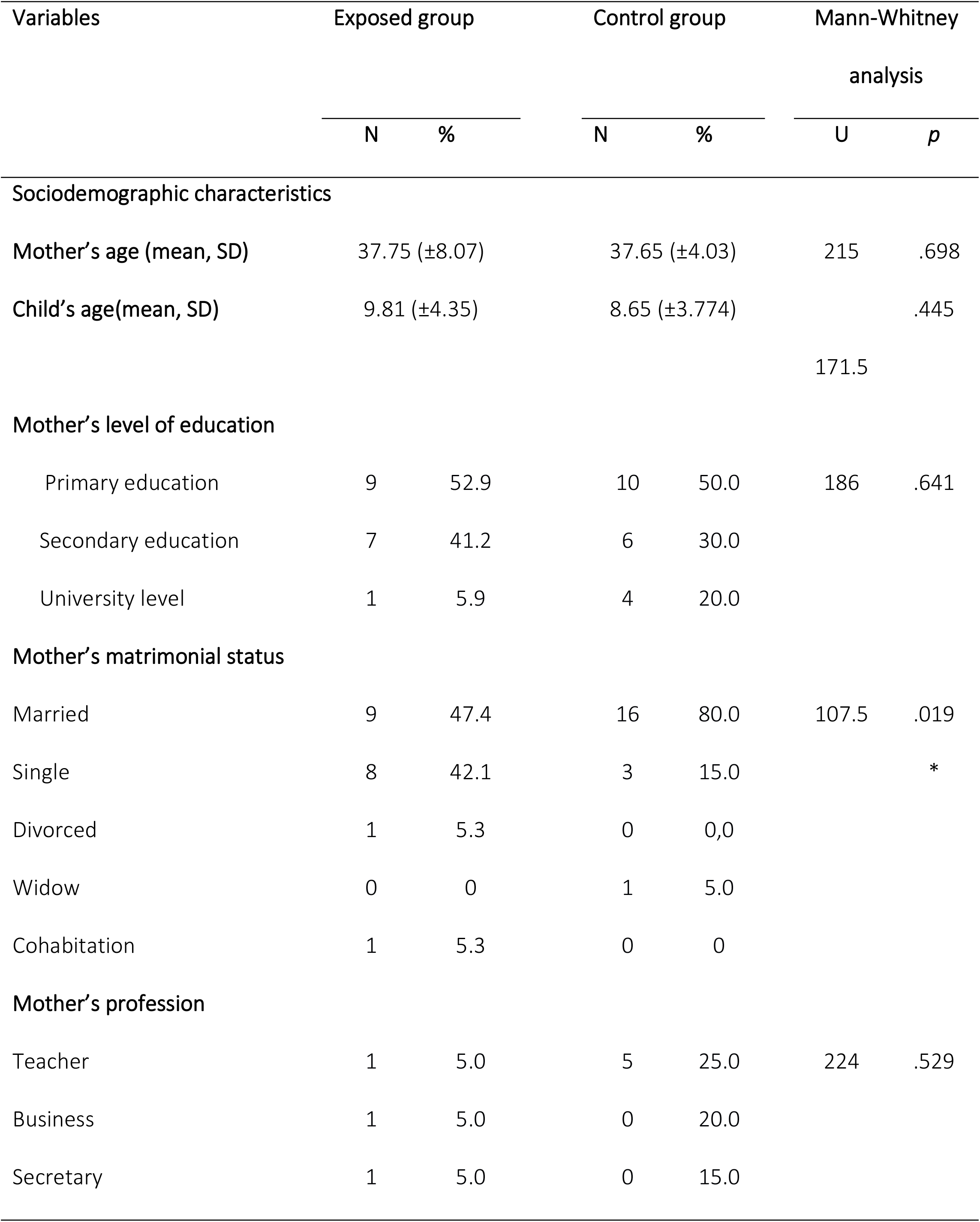
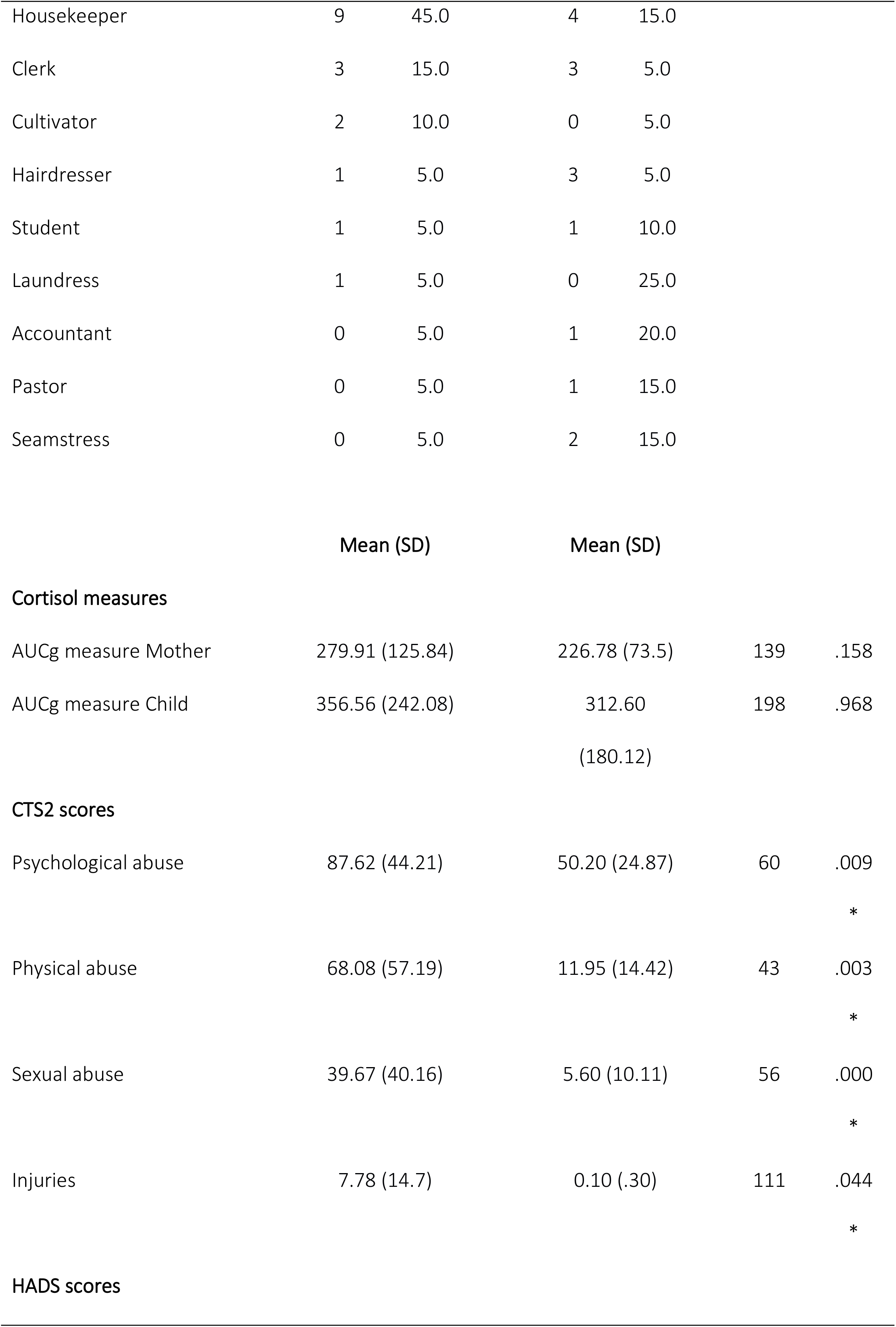
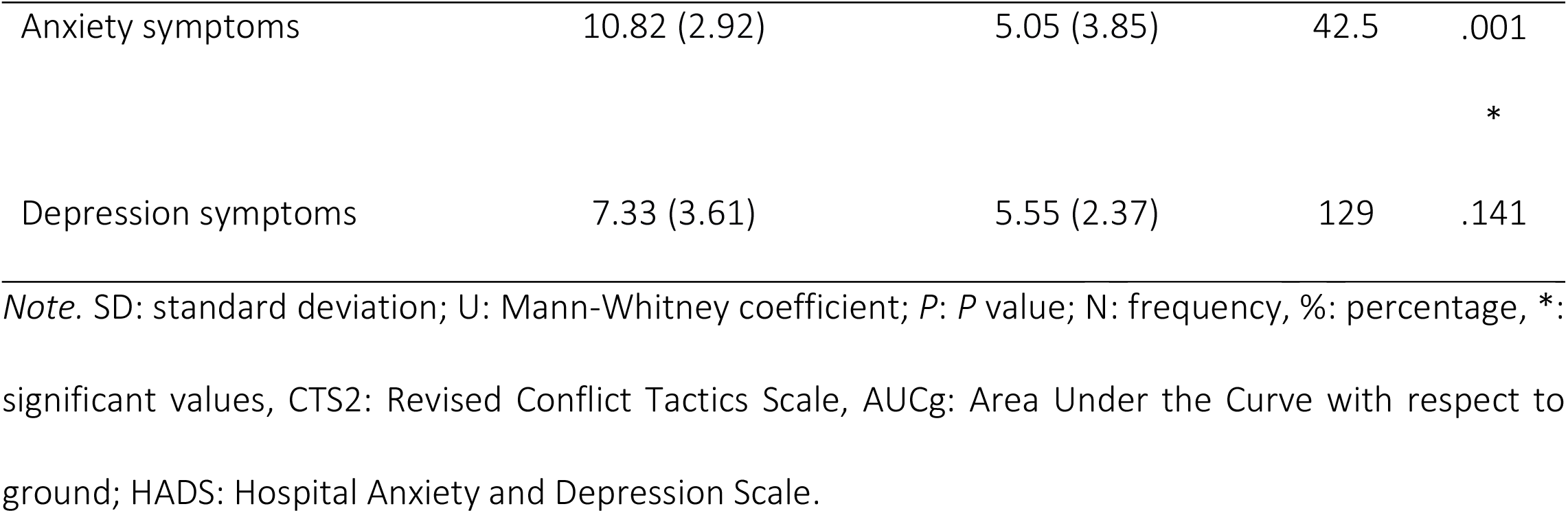
Sociodemographic characteristics, cortisol measures, CTS and HADS scores of participants

## Material and methods

After obtaining informed written consent as well as assent or written informed consent on behalf of minors, mothers completed self-report questionnaires and provided saliva using Oragenes kits (OG-600 Oragene) for epigenetic analysis. Salicaps (IBL International GmbH) were also used for salivary cortisol assessment. Each mother was given a pack containing six labeled Salicaps—three for themselves and three for their children—in which to collect saliva three times the next day at home: immediately after waking up, 30 minutes after waking up, and 45 minutes after waking up. Detailed information about the procedure for cortisol assessment can be found in an associated publication (17).

### Measures

#### Assessment of IPV

IPV was measured using the Revised Conflict Tactics Scale [CTS2; 37, 1996; French version by 38,1997]. This 78-item version explores violence between the mother and her partner in the past 12 months on five scales: negotiation skills (e.g. ‘I explained my side or suggested a compromise’), psychological abuse (e.g. ‘insulted, stomped out of room, or threatened to hit me’), physical abuse (e.g. ‘pushed, kicked, burned, scalded, or slapped me’), sexual abuse (e.g. ‘used force, used threats, to make me have sex’), and injuries (e.g. ‘felt pain, needed to see a doctor because of a fight’). The CTS2 is scored by adding the midpoints of the response categories chosen by the participant. The midpoints were 0 = 0 times, 1 = 1 time, 2 = 2 times, 4 = 3–5 times, 8 = 6–10 times, 15 = 11–20 times, and 25 = more than 20 times. Since there is no total score of the CTS2 and no validated cut-off score, we used the subscale scores, namely the scores for psychological abuse, physical abuse, sexual abuse, and injuries, and analyzed each of the subscales of the CTS2 separately. The global Cronbach’s alpha coefficient for this scale was 0.88. Looking at the subscales, we found a Cronbach’s alpha of .57 for psychological abuse, .71 for physical abuse, .50 for sexual abuse and .79 for injury.

#### Mother mental health indicators

Hospital Anxiety and Depression Scale [HADS; 39; French version by 40] was used to examine mothers’ current anxiety and depression symptoms. The HADS is composed of 14 items rated from zero to three, with seven questions related to anxiety and seven others related to depression. Two scores are obtained, with a minimum of zero and a maximum of 21. Higher scores indicate higher levels of anxiety and depression (Cronbach’s coefficient: anxiety subscale =.72 and depression subscale =.61).

#### Cortisol measures

The participants were instructed to bring the samples immediately after collection to the ALVF premises. In total, we received 240 samples for cortisol assessment. All samples (n = 240) were kept cool in a refrigerator for a few days in Yaoundé (Cameroon). Afterwards, the samples were sent to Fribourg (Switzerland), where they were frozen and stored at –20°C before being sent to Dresden (Germany) for analysis and assaying in the Laboratory of Biopsychology of the Technical University of Dresden, Germany (Luminescence Immunoassay, IBL). After thawing, the SaliCaps were centrifuged at 3,000 rpm for 5 min, which resulted in a clear supernatant of low viscosity. Salivary concentrations were measured using commercially available chemiluminescence immunoassay with high sensitivity (IBL International, Hamburg, Germany).

#### Saliva sampling for genomic DNA extractionn

Saliva was collected using the OG-600 Oragene DNA self-collection kit following the manufacturer’s instructions (DNA Genotek). Each participant was asked to provide 2 ml of saliva, which was mixed with 2 ml of the Oragene solution, beginning the initial stage of DNA isolation and stabilizing the sample until extraction could be performed. In total, we received 80 samples for DNA methylation analysis which, like cortisol samples, were kept cool in a refrigerator for a few days in Yaoundé and sent to Fribourg, where they were frozen and stored at -20°C.

#### Sodium bisulfite sequencing

Genomic DNA was isolated from the saliva of the 80 participants using the kit Saliva DNA Isolation 96-Well Kit from Norgen (RU35200). Genomic DNA bisulfite conversion was achieved using EpiTect Fast 96 DNA Bisulfite Kit from Qiagen. Amplicons of 406 bp from the 1F genomic region of the NR3C1 gene (chromosome V from 143’404’319 to 143’403’963 -GRCH38.p12 version of the genome) were generated by PCR as described in Perroud et al. 2011. Oligonucleotides used for the amplification were as followed: forward primer 5’-TTTGAAGTTTTTTTAGAGGG-3’ and reverse primer 5’-CCCCCAACTCCCCAAAAA-3’. Libraries for Illumina sequencing were prepared according to the manufacturer instructions. Amplicons were sequenced on an Illumina MySeq platform at the SICCH (Swiss Integrative Center for Human Health, Fribourg, Switzerland).

### Data analysis

Output sequences were processed to deliver percentages of methylation at each of the 47 CpG dinucleotides examined. The reads were quality controlled with FastQC (v0.11.9) (41) and MultiQC (v1.8.1) (42). Mapping of the raw reads to the region of interest (3936bp 5’-UTR of gene NR3C1 chrom5:143401879-143405816) was performed with bwa-meth (v0.2.2) (43) and to validate the data, some of the bam files were visualized with IGV (v2.8.13) (44). Counting the methylations positions at CpGs was carried with MethylExtract (v1.9.1) (45) leading to a tab-delimited file containing the methylated and unmethylated counts for each sample and each CpG position. Each of the 47 CpG positions within the region of interest was evaluated and transformed into percent methylation. To avoid sequencing quality issues, the counts were calculated from the best quality regions, meaning from the forward reads for the most 5’ CpG #1 (CpG pos 1495 to CpG pos 1757) and from the reverse reads to the last most 3’ CpG #47 (CpG pos 1771 to CpG pos 1850). Zero counts for unmethylated regions were transformed to “NA” to avoid calculation issues.

Methylation analysis was conducted regarding global mean methylation level and regarding specific CpG methylation level. Global *NR3C1* DNA methylation levels was computed as one mean score per individual indexing overall DNA methylation levels across all the CpG islands of the exon 1F. Based on previous research (33, 29), specific CpG sites in the promoter region proximal to a known transcription binding factor were identified as : CpG 12-13, CpG 16-21, CpG 30-32, CpG 37-38 and their DNA methylation level was calculated.

First, we tested the normality of the data including anxiety and depression measures, and cortisol and DNA methylation measures. Normal Q-Q plots and box plots showed that almost all variables were not normally distributed (all *p*_s_ < .05). The exceptions were the anxiety and depression scores on HADS. Comparison between groups were made using a non-parametric Mann-Whitney *U* test. Spearman correlation was used to test basic associations between DNA methylation and cortisol concentration respectively mental health scores.

To determine the total concentration of cortisol post awakening, we computed the cortisol area under the curve with respect to the ground (AUCg) according to the formula described by 46 (2013). Since cortisol and methylation data were skewed, we performed log-transformation of the data to achieve normal distribution before testing our models.

The associations between DNA methylation and cortisol concentration respectively symptoms of psychopathology were explored in linear regression models, using mother’s mental health scores as outcome variable and methylation level as the independent variable. The exposed group was coded as 0 and the control group was coded as 1. other covariates (such as cortisol level, age, profession, level of education) were added to the model as explanatory variables.

All analyses were conducted using SPSS for Windows, version 28.

## Results

A total of 40 dyads (mother and children) participated to the study. 20 dyads were exposed to IPV and 20 were used as a control group. Mothers were aged between 25 and 53 years and children between 2 and 18 years. Sociodemographic characteristics showed significant differences between the two groups on matrimonial status (U = 107.5, *p =*.019) using Mann-Whitney analysis. There were no significant group differences for mother’s profession, mother’s level of education, mother’s age and child’s age (all *p* >.445) (Table 1). IPV exposed mothers reported higher rates of different types of abuse. Analyses showed significant group differences on psychological abuse (U = 60, *p* = .009), physical abuse (U = 43, *p* = .003), sexual abuse (U = 56, *p* = .000), and injuries (U = 111, *p* = .044). Assessing mental health indicators (HADS score), we observed high levels of anxiety in the exposed group compared to the control group (U = 42.5, *p* < .001). No significant differences across the groups were found for depression. Altogether, the exposed group exhibited significantly higher level of various types of abuse associated with anxiety (see Table 1).

### Cortisol measures

In order to determine whether there are evidence of HPA axis deregulation, we measured the level of cortisol from saliva in the different groups. Although we observed a higher level of cortisol in exposed groups (mothers and children), our results indicated no significant difference between the exposed and the control groups, for mothers (U = 139, *p* = .158) and children (U = 198, *p* = .968).

### DNA methylation comparison between groups for mothers and children

We examined the methylation level of the 47 CpG dinucleotides in the exon 1F promoter region of the NR3C1 gene to assess whether there is a link between exposure to IPV and methylation of a gene involved in HPA axis regulation. Regarding the mothers, when comparing the average *NR3C1* methylation level of the 47 CpG island between the groups, our results showed a higher methylation level in the exposed group, however, the difference was not significant. Next, we examined in more details, 4 groups of CpGs sites that correspond to putative binding sites for the NGFI-A transcription factor, which is known to regulate transcription of the NR3C1 gene. We found a significantly higher methylation percentage at CpG 16-21 in the exposed group (U = 119,5, *p* = .028) (Table 2), relative to the methylation level in control group. The effect size for this analysis (d = .73) was medium according to Cohen’s (47) convention.

**Table 2.** Mean and site-specific DNA methylation comparison across groups of mothers and children

|  | Exposed group | Control group | Mann-Whitney analysis |  |
| --- | --- | --- | --- | --- |
|  | M (SD) | M (SD) | U | p |
| **Mothers** |  |  |  |  |
| Global mean methylation | 0.89 (0.28) | 0.76 (0.30) | 149 |  |
|  |  |  |  | .174 |
| **Site specific mean methylation** |  |  |  |  |
| CpG 12-13 | 0.79 ($\pm 0.76$) | 0.51 ($\pm 0.67$) | 160,5 | .258 |
| CpG 16-21 | 0.80 ($\pm 0.35$) | 0.47 ($\pm 0.53$) | | .028 |
|  |  |  | 119.5 | * |
| CpG 30-32 | 0.97 ($\pm 0.50$) | 1.11 ($\pm 0.66$) | 234 | .357 |
| CpG 37-38 | 0.09 ($\pm 0.24$) | 0.30 ($\pm 0.51$) | 236 | .183 |
| **Children** |  |  |  |  |
| Global mean methylation | 0.74 (0.40) | 0.74 (0.45) |  | 181 |
|  |  |  |  | .813 |
| **Site specific mean methylation** |  |  |  |  |
| CpG 12-13 | 0.59 ($\pm 0.75$) | 0.54 ($\pm 1.10$) | 165,5 | .496 |
| CpG 16-21 | 0.44 ($\pm 0.41$) | 0.63 ($\pm 1.12$) | 186 | .923 |
| CpG 30-32 | 1.05 ($\pm 0.92$) | 0.79 ($\pm 0.72$) | 160.5 | .411 |
| CpG 37-38 | 0.07 ($\pm 0.24$) | 0.08 ($\pm 0.35$) | 182 | .835 |

**Note.** U= Mann-Whitney coefficient, M (SD) = mean (standard deviation), p= p-value, \*= significant values

The same methylation analysis was performed with children. However, there was no significant difference in global mean methylation and methylation level at NGFI-A binding sites between the groups.

### Association between DNA methylation and mother’s anxiety and depression symptoms

Spearman rho correlation analysis showed in the exposed group as compared to the control group, a significant positive association between the level of methylation at CpG 16_21 and symptoms of anxiety (r = .507, *p* = .038) (see Fig. 2). The association with depression symptoms did not reach significance.

**Fig 2.**
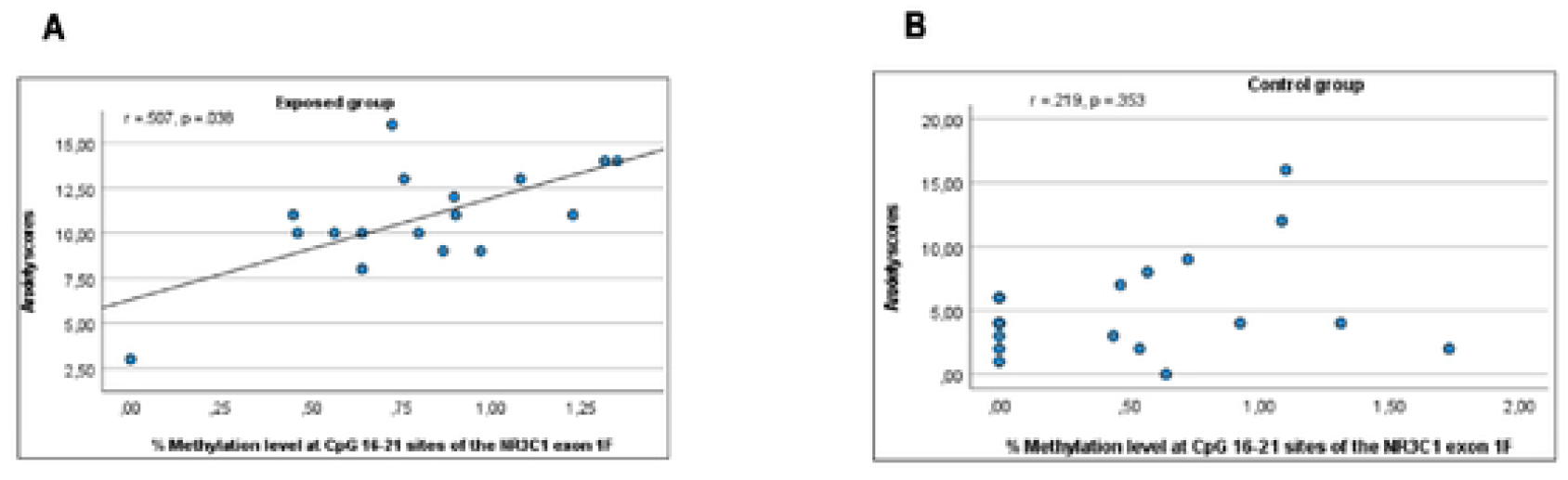
Scatter Plot of anxiety scores by methylation level at CpG 16-21 sites of the NR3C1 exon lF. **A)** represent significant results obtained in the exposed group (r = .507, *p* = .038) and **B)** showed non-significant results obtained in thecontrol group (r= .219, *p* = .353).

Further linear regression model testing the effect of methylation level on mother’s anxiety was statistically significant, F(6) = 6.495, *p* < .001. The model explained 59% of the variance. Parameter estimate showed statistically significant results for group (B = -4.931, *p* < .001, 95% CI, -7.485 to -2.377) and methylation level at CpG 16_21 (B= 2.614, *p* = .046; 95% CI, 0.047 to 5.182). Other parameter estimate such as age, profession, level of education and cortisol level were not significant (all *P* values >.202).

Taken together, our results showed a significant positive association between the level of methylation at CpG 16-21 sites and symptoms of anxiety in the exposed group as compared with the control group.

## Discussion

This study on IPV in sub-Saharan Africa provides evidence for epigenetic modifications associated with IPV known to be highly prevalent in this region. The most interesting result of this study was the significant difference in methylation level at CpG 16-21 sites of the *NR3C1* exon 1F promoter region in the exposed mothers as compared to the control group. This CpG group encompasses a putative NGFI-A binding site, that might be involved in the regulation of NR3C1 gene transcription. In addition, there was a significant positive association between the level of methylation at CpG 16-21 sites and mother’s mental health in particular anxiety symptoms in the exposed mothers group compared to the control group.

The results are consistent with our hypothesis. The differential methylation level at CpG 16-21 sites which corresponds to a NGFI-A putative binding site, suggests that GR gene expression in exposed mothers might be down-regulated. However, given that the observed increase in methylation level was subtle, the deregulation of the HPA axis may not be strong. Moreover, this interpretation is consistent with the absence of a significant correlation between methylation level and cortisol concentration in mothers and their children exposed to IPV.

Concerning the children, our results showed no significant DNA methylation difference between the groups. These results are in contrast to the findings of 34, who found that methylation status of the GR gene of adolescent children was influenced by their mother’s experience of IPV. A possible explanation for such discrepancies can lie in the type of tissue used and the developmental period studied. While 34 studied the effect of IPV before birth and used blood samples, we investigated the postnatal period (with children aged 2 to 18 years) and examined saliva samples which may show lower levels of methylation compared to blood DNA methylation as evidenced by 48. Furthermore, the small sample size may greatly limit the statistical power to detect the relationship as statistically significant.

One striking finding was the very low level of methylation at the canonical NGFI-A binding site (CpGs 37-38) in all groups. 1F CpG #37 is the most widely studied CpG in both animal and human. Higher level of methylation has been associated with a wide variety of early life stress (reviewed in 26, 33). However, there are also studies that report decreased level of methylation at this particular CpG in response to individuals with PTSD or in newborns who were exposed to maternal smoking or maternal anxious-depressed condition (reviewed in 26). One possible interpretation of our results is that the NR3C1 receptor gene is still actively regulated, at least in the cells included in the saliva samples. This hypothesis is in line with our cortisol measurements, which do not point towards a strong deregulation of the HPA axis.

Exposed mothers in this study have different forms of abuse (e.g. injuries, psychological, physical and sexual abuse) and also differ in anxiety and depression symptoms. Within the exposed group, we found a significant positive association between symptoms of the mothers in anxiety and level of methylation at CpG 16-21 sites. This suggest that methylation of the CpG 16-21 sites, though average, may have the potential to induce changes in the HPA axis function by increasing the risk of poor mental health. CpG 16-21 methylation might confer a certain vulnerability. These results add to accumulating knowledge of the mechanisms by which early adversity in particular IPV contributes to developmental trajectories and psychopathology in IPV victims.

### Limitations

When interpreting the results of the present study, several limitations need to be taken into account. First, methylation levels were low in mothers and even lower in children (less than 10%). Although the observed differences in DNA methylation between the exposed and control group were statistically significant at the CpG 16-21 sites, they were medium effect sizes. Second, the sample size was small limiting the power of the conclusion. Third, epigenetic analysis was based on saliva samples, which can account for small level of methylation as compared to blood sample as shown in previous studies (48). Perhaps, saliva might not be the ideal proxy for methylation measurements as suggested by a previous finding (49). Fourth, we used the conflict tactic scale (CTS2) which is a self-reported retrospective instrument used to measure IPV with some subscales like psychological abuse showing low reliability (.57). Fifth, salivary cortisol which is prone to substantial intra-individual variability and limited reliability (50), was examined. Lastly, for cortisol assessment, we have three measurements per person on a single day which can lead to significant negative impact on estimate of the total cortisol concentration. However, in this context, many challenges prevented us from collecting saliva for more than one day. First participants negatively perceived giving their saliva and second during data collection, some women were banned by their partners to participate again in the study.

Despite these limitations, this research has laid the basis for identifying critical links between psychological parameters and biological markers, such as cortisol levels and *NR3C1* gene methylation in mother-child dyads exposed to domestic violence.

### Clinical implication

This is the first study reporting the link between IPV and DNA methylation of *NR3C1* exon 1F promoter region in Sub-Saharan Africa. We provide evidence of epigenetic alterations associated with IPV in a non-western cultural setting where numerous social norms, are known major risk factors for IPV (51, 52).

The results of this study have several clinical implications for women mental health. Firstly, this study highlighted that IPV is detrimental to the well-being of women exposed to it. This result is strengthened by the inclusion of methylation and cortisol analyses from saliva samples as well as mental health measures. There is an urgent need to create awareness on this. In order to reach all levels of society, particularly in sub-Saharan Africa, community workers may serve as a key resource due to their networking with the local population, NGOs, and IPV professionals. Secondly, we identified a NGFI-A putative binding site (CpG 16-21) that is more methylated in exposed mothers and which may have the potential to confer vulnerability for psychopathologies. This evidence may assist professionals with timely prevention and intervention strategies

## Conclusion

Our result showed a significant difference in DNA methylation at CpG 16-21 sites of the *NR3C1* proximal promoter (exon 1F) between the exposed and control group. This study lays the foundation for future epigenetic studies in sub-Saharan Africa. Still, our results warrant replication. Future studies with ethnically diverse samples can provide more robust conclusions. Epigenetic methylation of related stress genes such as the *FK506* binding protein 5 (*FKBP5*) gene and the oxytocin receptor (*OXTR*) gene could provide further insights into potentially more pronounced impact of IPV and their role in stress-related disorders.

## Data Availability

All data files are available from the Inter-university Consortium for Political and Social Research (ICPSR) database (DOI: https://doi.org/10.3886/E177242V1)

## Acknowledgements

We are thankful to the families who participated and contributed to this work, as well as to the ALVF in Cameroon and their community agents for facilitating the recruitment of participants. We would also like to acknowledge Prof. Ketcha Wanda with whom we started this project and who passed away. Finally, we thank Prof. Linda Booij for her support and valuable feedback.

## Data reporting

The data that support the findings of this study are openly available in https://doi.org/10.3886/E177242V1

## Authors’ contributions

**DLW, NM, CMS** and **CW** designed and directed the project; **DLW** collected and analyzed the data; **CW** performed the epigenetic analysis; **DLW** wrote the first draft of the manuscript which was reviewed and edited by **CW** and **CMS**.

